# Glucagon-like peptide-1 receptor agonist-induced lipidome remodelling is associated with improved liver, kidney and inflammatory markers in type 2 diabetes

**DOI:** 10.64898/2026.07.22.26358565

**Authors:** Daria Lipska, Tommi Suvitaival, Simon Maria Kienle, Bernt Johan von Scholten, Rasmus Sejersten Ripa, Emilie Hein Zobel, Joachim Størling, Martin Bæk Blond, Tarunveer S. Ahluwalia, Tine Willum Hansen, Lotte Bjerre Knudsen, Mads Almose Røpke, Joana Mendes Lopes de Melo, Karolina Sulek, Cristina Legido-Quigley, Peter Rossing

**Author notes:** Corresponding authors: Karolina Sulek, Cristina Legido-Quigley, and Peter Rossing. These authors contributed equally (Daria Lipska, Tommi Suvitaival, Simon Maria Kienle).

## Abstract

**Introduction:** Lipids are considered both drivers and biomarkers of cardiometabolic diseases. As glucagon-like peptide-1 receptor agonists (GLP-1RAs) are widely used for diabetes and obesity management, it is crucial to understand how they affect the related comorbidities through the circulating lipidome. This study investigated the lipidomic changes induced by liraglutide treatment when compared to placebo in people with type 2 diabetes (T2D) to characterise lipid remodelling and its association with clinical outcomes.

**Research design and methods:** This post-hoc study analysed plasma samples using liquid chromatography-mass spectrometry (LC-MS/MS) from LIRAFLAME, a randomised, double-blind, placebo-controlled, parallel-group trial. A hundred people with T2D received up to 1.8 mg of liraglutide or placebo once daily for 26 weeks. Plasma samples were collected at baseline, week 13 and week 26.

**Results:** Liraglutide treatment resulted in a statistically significant increase in multiple lysophospholipid subclasses, including LPCs, LPC(O)s, LPC(P)s, LPEs, and LPE(P)s, observed at 13 weeks and sustained at 26 weeks compared to placebo. These increases were not mediated by the change in BMI. Triglyceride concentrations decreased at 13 weeks, while fatty acid levels declined at 26 weeks, consistent with enhanced lipid remodelling. The increase in LPC(O)s was associated with favourable decreases in ALAT, MCP-1, and UACR, suggesting anti-inflammatory effects with hepatic, renal, and cardiovascular benefits.

**Conclusions:** Compared to placebo, 26 weeks of liraglutide treatment resulted in a favourable lipidomic shift from a triglyceride-rich profile towards one enriched in lysophospholipids. This lipid remodelling was associated with improvements in hepatic, renal, and inflammatory markers.

**Graphical abstract:** 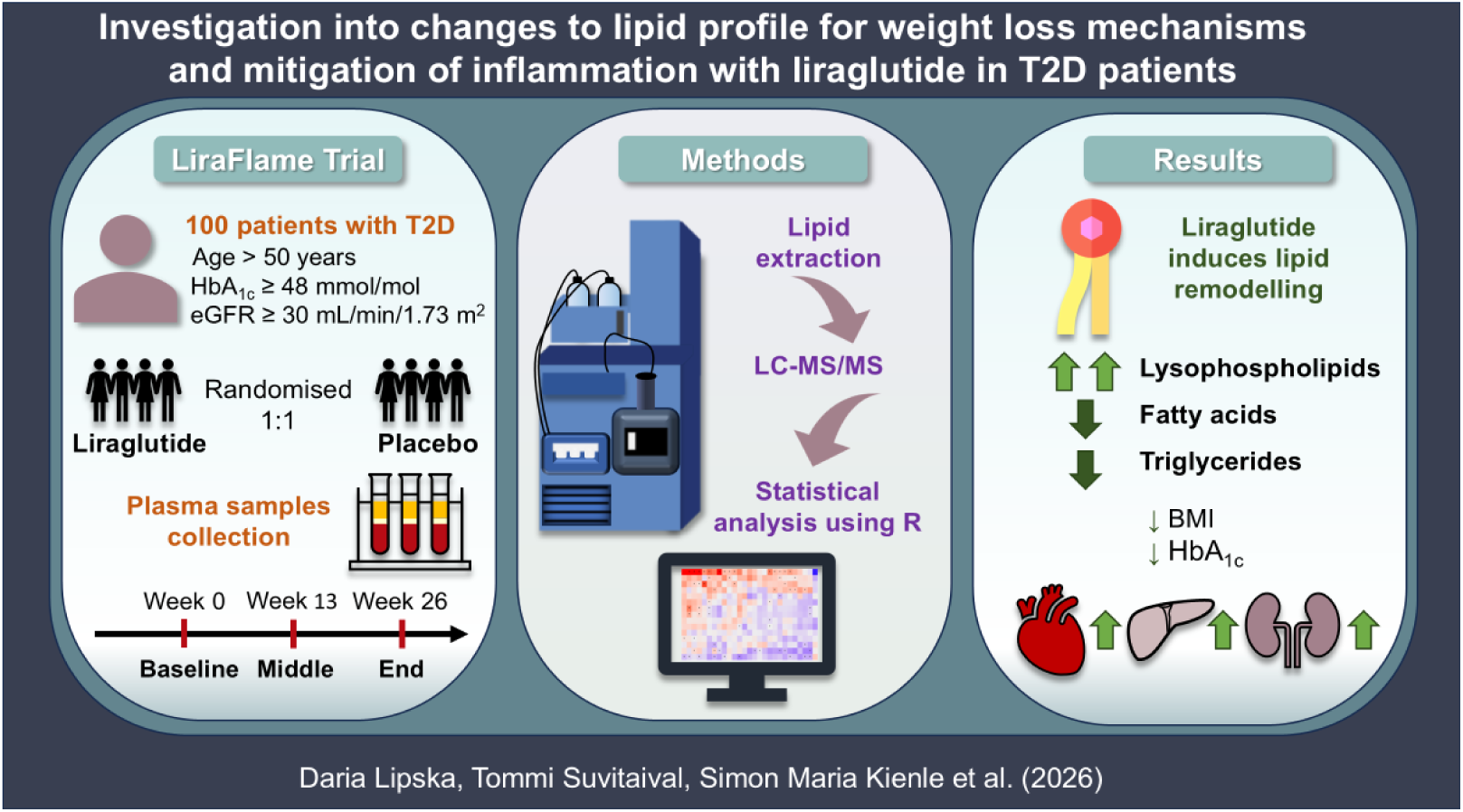

**Highlights:**

- Liraglutide upregulates lysophospholipids and lowers triglycerides and fatty acids.
- The strongest increase was in lysoalkylphosphatidylcholines (LPC(O)s).
- Lysophosphatidylcholines (LPCs) increased independently of weight loss.
- LPC(O/P)s associated with improvements in liver, kidney, and inflammation markers.

## 1. INTRODUCTION

Type 2 diabetes (T2D) is a chronic metabolic disorder that accounts for approximately 96% of all diabetes cases. The global prevalence of T2D is continuing to rise due to a poor diet and sedentary lifestyle.[1] The latest estimates from the International Diabetes Federation indicate that diabetes affects 1 in 9 adults worldwide.[2] T2D is characterised not only by chronic hyperglycaemia but also by broad cardiometabolic comorbidities, including cardiovascular disease (CVD), stroke, chronic kidney disease (CKD) and MASH.[3,4] Among the most relevant pathophysiological processes driving these complications, are dysregulated lipid metabolism, low-grade inflammation, and impaired metabolic and vascular signalling, which may affect the liver, kidney, and the vascular system. Accordingly, therapeutic approaches that improve glycaemic control and promote weight loss are also expected to beneficially modulate inflammatory and tissue-specific lipid-biomarkers. However, the molecular mechanisms linking treatment response to downstream clinical effects remain incompletely characterised [5].

Glucagon-like peptide-1 receptor agonists (GLP-1RAs), including liraglutide, improve glycaemic control and promote weight loss in people with T2D.[6,7] In addition to these metabolic benefits, liraglutide reduces the risk of major adverse cardiovascular events, cardiovascular death, and all-cause mortality in individuals with T2D at high cardiovascular risk.[8] GLP-1RAs also improve several cardiometabolic risk factors, including systolic blood pressure and fasting triglyceride concentrations, potentially through effects on inflammation, endothelial function, and tissue metabolism.[9,10] Translating these clinical observations into actionable mechanistic insight requires a deeper characterisation of systemic metabolic changes during treatment.

Lipids play fundamental roles in metabolic homeostasis, functioning as structural components of biological membranes, key mediators of signalling pathways, and regulators of energy balance and inflammation.[11,12] Specific lipid subclasses and molecular species have distinct biological roles, such that class-level increases may represent different mechanistic processes depending on chain length, saturation, and oxidation state [13]. Thus, time-resolved lipid profiling may identify pharmacodynamic signatures of liraglutide and help explain how systemic lipid pathway changes relate to clinical improvement. Lipidomics, with a high-resolution mass spectrometry approach, provides an opportunity to capture treatment-responsive lipid pathways. Various studies have reported individual lipids as critical signals for T2D and its complications.[14–19] In our previous work, daily liraglutide injections were shown to improve lipid profiles and downregulate triglycerides, phospholipids, and ceramides, while also reducing the risk of CVD.[9,20]

In this post-hoc analysis, we report the lipidomic changes induced by liraglutide treatment in people with T2D in the LIRAFLAME trial using targeted semiquantitative liquid chromatography-mass spectrometry (LC-MS/MS) analysis on plasma samples from three measurement points: baseline, after 13 and 26 weeks of treatment. We aimed to investigate the temporal pattern of these changes to elucidate the mechanisms by which weight loss and glycaemic control influence treatment outcomes and the mitigation of related comorbidities. Hence, we examined the association between lipid classes and inflammatory markers, as well as cardiovascular, hepatic, and renal health biomarkers, to provide further insight into the lipid-driven benefit of GLP-1RAs in people living with T2D.

## 2. MATERIALS AND METHODS

### 2.1 Clinical Trial

Plasma samples were collected during the clinical trial “Effect of Liraglutide on Vascular Inflammation in Type-2 Diabetes” (LIRAFLAME), registered at ClinicalTrials.gov with the trial registration number NCT03449654, previously described.[9,20–22] In brief, 100 individuals with T2D were randomised in a double-blind, placebo-controlled, parallel-group trial. Participants were treated with up to 1.8 mg of liraglutide or a placebo once daily for 26 weeks. The eligibility criteria were T2D, age > 50 years, glycated haemoglobin A_1c_ (HbA_1c_) ≥ 48 mmol/mol, and estimated glomerular filtration rate (eGFR) ≥ 30 mL/min/1.73 m^2^. The plasma samples were collected at three visits: at the start (visit 0, baseline), middle (visit 3 at week 13), and end of the trial (visit 5 at week 26) and stored at -80°C. Plasma samples were collected from fasting participants with a few exceptions in visit 3.

### 2.2 Lipid Extraction

Lipid extraction was performed as described previously.[23] In brief, 10 µL of plasma was mixed with 100 µL of butanol:methanol (1:1) with 10 mM ammonium formate, which contained a mixture of internal standards (Supplementary Methods). Samples were vortexed thoroughly and set in a sonicator bath for 15 min, maintained at room temperature, and subsequently incubated for 45 min at 1000 rpm at 4°C. Samples were then centrifuged (14,000 x g, 10 min, 4°C) before transferring into sample vials with glass inserts. Samples were stored at -80°C until analysis.

### 2.3 Liquid Chromatography – Mass Spectrometry

Analysis of plasma extracts was performed on an Agilent 6460C QQQ mass spectrometer with an Agilent 1290 series HPLC system and an ACQUITY UPLC CSH C18 column (2.1 x 100 mm, 1.7 µm, Waters) with the thermostat set at 45°C. Mass spectrometry analysis was performed by positive/negative ion mode switching with dynamic scheduled multiple reaction monitoring (dMRM). Mass spectrometry settings and MRM transitions for each lipid class, subclass, and individual species are shown in the Supplementary Methods.

The LC buffer consisted of solvent A: 50% H₂O / 30% acetonitrile / 20% isopropanol (v/v/v), containing 10 mM ammonium formate and 2 mM ammonium fluoride; and solvent B: 1% H₂O / 9% acetonitrile / 90% isopropanol (v/v/v) containing 10 mM ammonium formate and 2 mM ammonium fluoride. We used a stepped linear gradient with a 16- minute cycle time per sample and a 5 µL sample injection. The gradient detail can be found in the Supplementary Methods.

The following mass spectrometer conditions were used: gas temperature, 325°C; gas flow rate, 9 L/min; nebulizer, 35 psi; sheath gas temperature, 350°C; capillary voltage, 3000 V (both positive and negative); and sheath gas flow, 11 L/min. Isolation widths for Q1 and Q3 were set to ‘‘unit’’ resolution (0.7 amu).

### 2.4 LC-MS/MS Data Analysis and Lipid Quantification

Chromatographic peaks were integrated using the Mass Hunter (B.07.00, Agilent Technologies) software and assigned to a specific lipid species based on MRM (precursor/product) ion pairs and their chromatographic behaviour. To quantify lipid analytes, the ratio of each analyte peak with the corresponding internal standard was used.

### 2.5 Statistical Analysis

All data were analysed in R (version 4.4.1). Variables of the lipidomics data were standardized according to the variable-specific mean and standard deviation (SD) of the baseline time point. The reported treatment effects, therefore, are in population SD units.

The lipidomics data were primarily analysed according to lipid class and secondarily according to molecular species. All treatment effects were estimated with the longitudinal analysis of covariance (ANCOVA) models that included the baseline value of the outcome as a covariate, accounting for between-individual differences at baseline. Therefore, the reported estimates represent causal estimates of the treatment effects relative to placebo rather than within-group changes over time. The model was calculated with and without adjustment for the baseline values of the covariates age, sex, body mass index (BMI), systolic blood pressure (SBP), HbA_1c_, high-density lipoprotein (HDL), low-density lipoprotein (LDL), very low-density lipoprotein (VLDL), total cholesterol, total triglycerides (TGs) and eGFR. Analyses according to lipid class were calculated with class-specific linear mixed effects models, where the molecular lipid species identity entered the model as a random intercept.

The mixed-effects models were calculated using the package “lmerTest”.[24] Analyses of single variables – i.e., clinical variables and molecular lipid species – were calculated with least-squares regression. P-values from the analyses of treatment effects on lipid classes and clinical measurements were corrected for multiple testing with the Benjamini-Hochberg method and are denoted as “p_BH_”. P-values from the secondary analysis of treatment effects on molecular lipid species were reported as is and are denoted as “p”. The threshold of statistical significance was set to 0.05.

Treatment effects on classes of lipids in the two follow-up time points were visualized as two forest plots, using the package “ggplot2”. Treatment effects on molecular lipid species were visualized as forest plots and as lipidome-wide heatmaps using the package “lipidomeR”.[25]

Two types of correlation analyses were calculated between all class sums of molecular lipid levels and those clinical measurements with any statistically significant treatment effect over the trial: First, correlation between pairs of change scores was calculated with a linear mixed effect model, including both the follow-up time points, and adjusting for treatment group. Second, cross-sectional Pearson correlation was calculated between lipid class totals and clinical measurements at baseline.

Finally, mediation analysis between the most affected lipid class and the affected clinical measurements was calculated using the R-package “mediate”.[26] Mediation analysis with three time points was calculated, where it was tested whether the treatment effect on the mediator at 13 weeks mediates the treatment effect on the outcome at 26 weeks, both adjusted for the baseline levels of the mediator and the outcome. Mediation analysis was calculated as lipid levels mediating clinical measurements as well as the reverse.

## 3. RESULTS

### 3.1 Baseline characteristics

The study analysed plasma samples collected from 100 people with T2D at three time points: baseline, mid-treatment (week 13), and end of treatment (week 26). The average age of the participants was 67 and 66 years in the placebo and treatment groups, respectively (Table 1). BMI was 29.2 kg/m^2^ in the placebo and 30.6 kg/m^2^ in the treatment groups. The largest baseline differences between groups were observed for total TGs: 1.56 mmol/L for the placebo and 2.08 for the liraglutide groups and alanine aminotransferase: 45 IU/L in the placebo and 40 in the treatment.

**Table 1.**
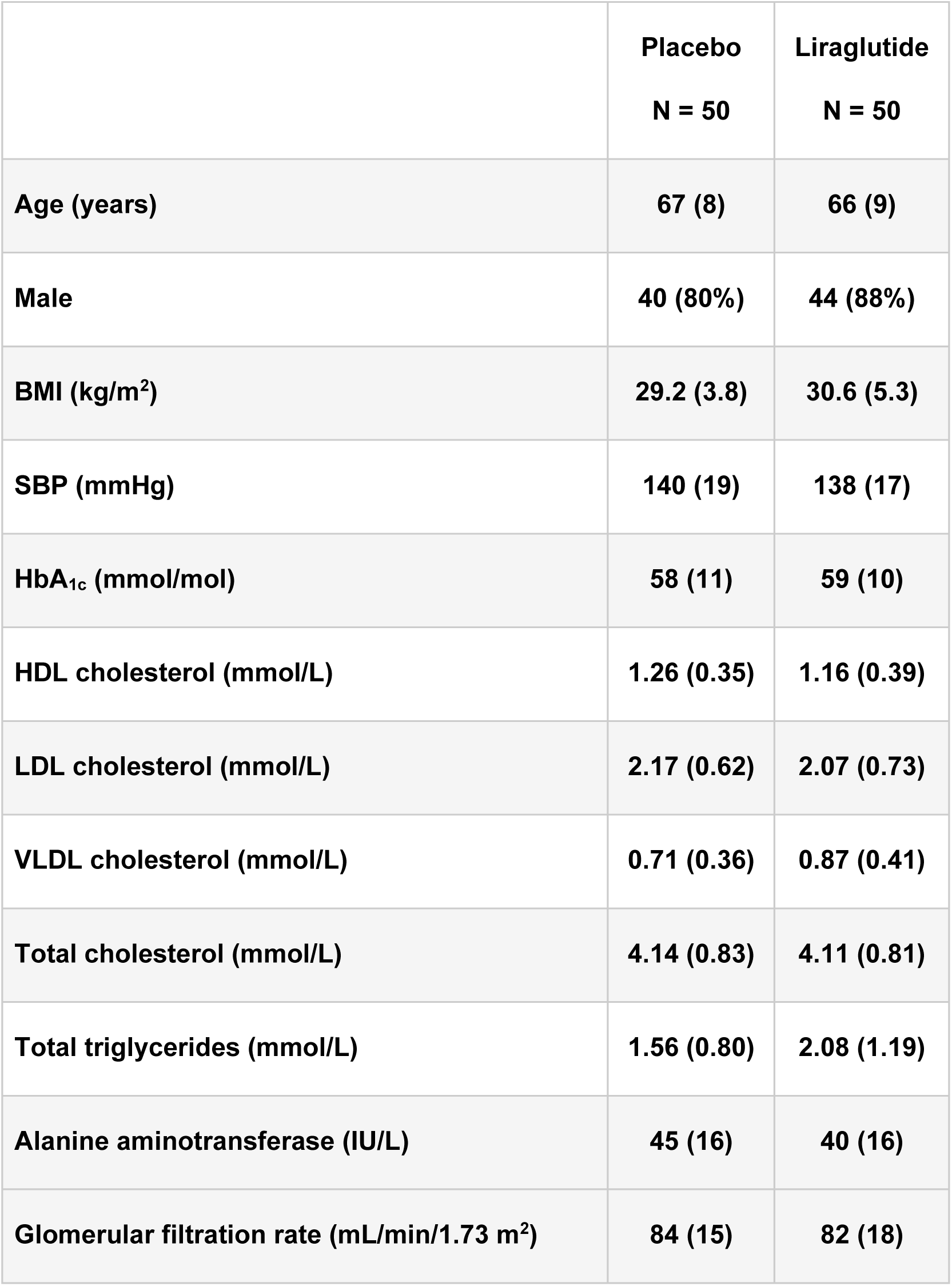
Clinical characteristics of the participants at baseline presented as mean (SD) or n (%).

|  | **Placebo**<br>**N = 50** | **Liraglutide**<br>**N = 50** |
| --- | --- | --- |
| **Age (years)** | **67 (8)** | **66 (9)** |
| **Male** | **40 (80%)** | **44 (88%)** |
| **BMI (kg/m<sup>2</sup>)** | **29.2 (3.8)** | **30.6 (5.3)** |
| **SBP (mmHg)** | **140 (19)** | **138 (17)** |
| **HbA<sub>1c</sub> (mmol/mol)** | **58 (11)** | **59 (10)** |
| **HDL cholesterol (mmol/L)** | **1.26 (0.35)** | **1.16 (0.39)** |
| **LDL cholesterol (mmol/L)** | **2.17 (0.62)** | **2.07 (0.73)** |
| **VLDL cholesterol (mmol/L)** | **0.71 (0.36)** | **0.87 (0.41)** |
| **Total cholesterol (mmol/L)** | **4.14 (0.83)** | **4.11 (0.81)** |
| **Total triglycerides (mmol/L)** | **1.56 (0.80)** | **2.08 (1.19)** |
| **Alanine aminotransferase (IU/L)** | **45 (16)** | **40 (16)** |
| **Glomerular filtration rate (mL/min/1.73 m<sup>2</sup>)** | **84 (15)** | **82 (18)** |

### 3.2 The effect of liraglutide on clinical measurements

Liraglutide was effective at lowering BMI and HbA_1c_ after 26 weeks of treatment (Supplementary Table 1), which has been previously reported (Ripa et al., 2021; Zobel et al., 2021). The average treatment effect for BMI was -1.01 kg/m^2^ (p=2.4×10^-15^) at 13 weeks and -1.04 kg/m^2^ (p=3.1×10^-16^) at 26 weeks, and for HbA_1c_, it was -6.59 mmol/mol (p=4.1×10^-9^) at 13 weeks and -4.90 mmol/mol (p=1.1×10^-5^) at 26 weeks. Total TGs showed a decrease of -0.30 mmol/L (p=0.019) at 13 weeks. There was no statistically significant effect measured for HDL, LDL, VLDL and total cholesterol. We observed a decrease in alanine aminotransaminase (ALAT) of -2.62 IU/L (p=0.12) at 13 weeks and -7.17 IU/L (p=2.4×10^-5^) at 26 weeks.

Moreover, we evaluated inflammatory, cardiac, and renal biomarkers to examine the effectiveness of liraglutide in mitigating diabetes-related complications (Figure 1A). At 13 weeks, pro-B-type natriuretic peptide (proBNP) and monocyte chemoattractant protein-1 (MCP-1) were decreased (-0.20 SD-units; Benjamini-Hochberg-corrected p-value, pBH=4.7e-4 and -0.24, pBH=0.010, respectively), whereas blood creatinine, thrombocytes, monocytes, and lymphocytes were increased (0.18, pBH=0.024; 0.24, pBH=0.0072; 0.4, pBH=0.010; 0.16, pBH=0.024; respectively). At 26 weeks, eGFR, urinary albumin-to-creatinine ratio (UACR), urinary albumin and intercellular adhesion molecule-1 (ICAM) were decreased (-0.19, pBH=0.014; -0.025, pBH=0.018; -0.05, pBH=0.041; -0.32, pBH=0.041; respectively), whereas blood creatinine as well as thrombocytes, and neutrophils were increased (0.22, pBH=0.0076; 0.19, pBH=0.037; 0.35, pBH=0.041; respectively).

**Figure 1.**
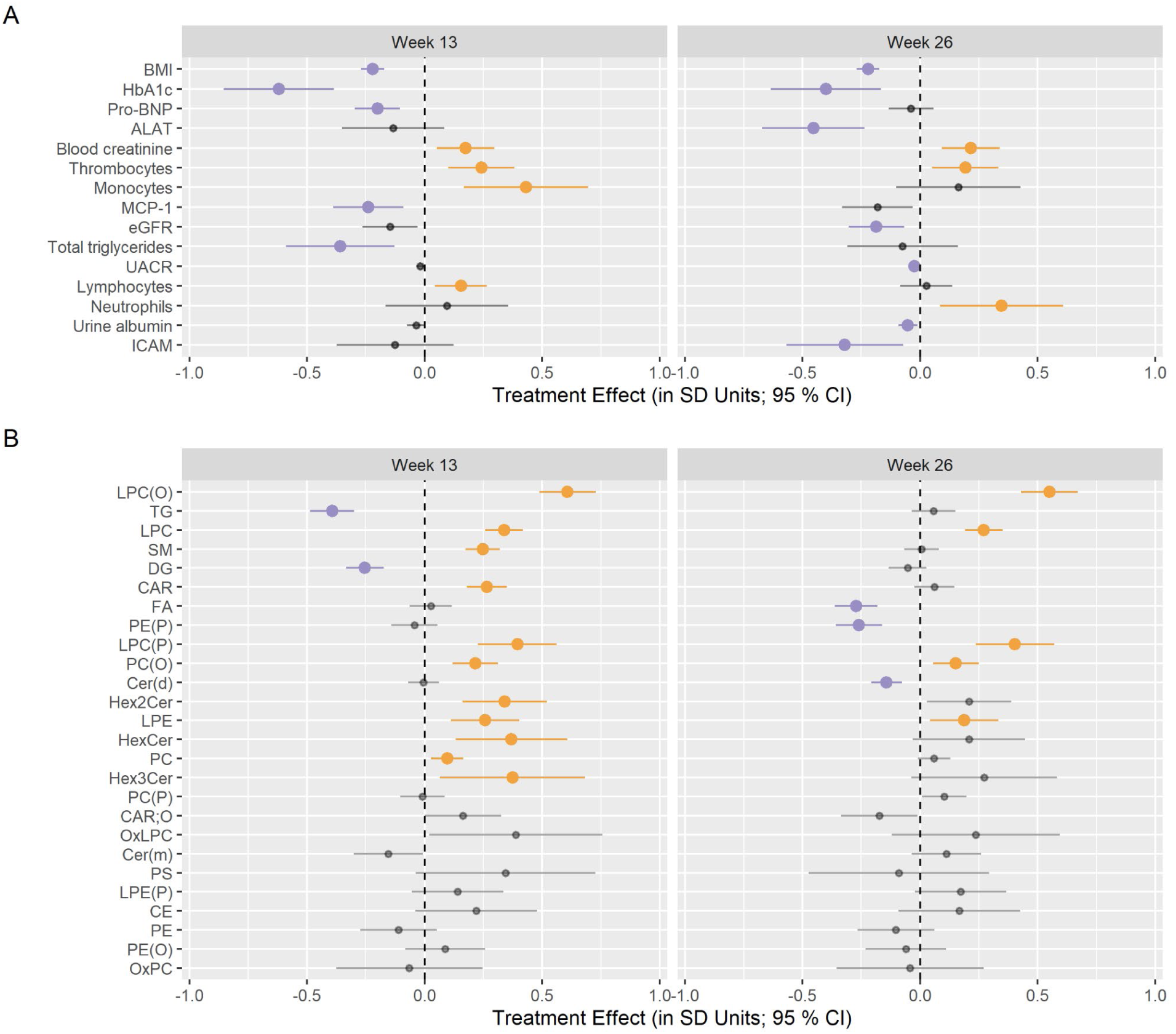
The effect of liraglutide treatment on the levels in A) clinical variables and B) lipid classes with adjustment to the baseline levels of the clinical covariates age, BMI, sex, HbA_1c_, ALAT, SBP, HDL, LDL, VLDL, total cholesterol, total TGs, and eGFR. The effects after 13 weeks (left) and 26 weeks (right) of treatment are shown. Increases and decreases with statistical significance (pBH<0.05) are highlighted in orange and purple, respectively.

### 3.3 The effect of liraglutide on plasma lipidome

A total of 513 lipids were measured in plasma, from the following lipid categories: glycerolipids, glycerophospholipids, sphingolipids, sterols, and fatty acyls (Figure 1B). The independent effect of liraglutide treatment on lipid class levels was confirmed in models with adjustments to the baseline levels of the clinical covariates age, BMI, sex, HbA_1c_, ALAT, SBP, HDL, LDL, VLDL, total cholesterol, total triglycerides (TGs), and eGFR. The changes in lipid classes before these are shown in Supplementary Figure 1.

Four subclasses of lysophospholipids: lysophosphatidylcholines (LPCs), lysoalkylphosphatidylcholines (LPC(O)s), lysoalkenylphosphatidylcholines (LPC(P)s) and lysophosphatidylethanolamines (LPEs) increased in level at 13 weeks and maintained the increase at 26 weeks: LPCs increased 0.34 SD-units (95% CI: 0.26, 0.42 SD; pBH=2.2e-15) at 13 weeks and 0.27 SD (0.19, 0.35; pBH=5.4e-10) at 26 weeks; LPC(O)s increased 0.61 (0.49, 0.73; pBH=4.0e-21) at 13 weeks and 0.55 (0.43, 0.67; pBH=2.2e-17) at 26 weeks; LPC(P)s increased 0.40 (0.23, 0.56; pBH=1.6e-5) at 13 weeks and 0.40 (0.24, 0.57; pBH=1.4e-5) at 26 weeks; and LPEs increased 0.26 (0.11, 0.40; pBH=1.4e-3) at 13 weeks and 0.19 (0.04, 0.33; pBH=0.037) at 26 weeks.

Alkenylphosphatidylcholines (PC(O)s) increased in level with 0.22 (0.12, 0.31; pBH=3.9e- 5) at 13 weeks and with 0.15 (0.06, 0.25; pBH=0.0079) at 26 weeks. Phosphatidylcholines (PCs) increased with 0.10 (0.03, 0,17; pBH=0.013) at 13 weeks and returned towards baseline level at 26 weeks (0.06; -0.01, 0.13; pBH=0.16).

Alkenylphosphatidylethanolamines (PE(P)s) were unchanged at week 13 (-0.04; -0.14, 0.06; pBH=0.46) but, in contrast to all other phospholipids, PE(P)s decreased with -0.26 (-0.36, -0.16; pBH=1.4e-6) at week 26.

Glycerolipids decreased at week 13: triacylglycerols (TGs) with -0.39 (-0.49, -0.30; pBH=1.4e-15) and diacylglycerols (DGs) with -0.25 (-0.33, -0.17; pBH=3.0e-9). These decreases were transient, and at 26 weeks TGs and DGs were unchanged from baseline (0.06; -0.04, 0.15; pBH=0.27, and -0.05; -0.13, 0.03; pBH=0.27, respectively). Inversely to glycerolipids, fatty acids (FAs) were unchanged at 13 weeks (0.03; -0.06, 0.12; pBH=0.64) but decreased in level with -0.27 (-0.36, -0.18; pBH=3.8e-8) at 26 weeks.

Acylcarnitines (CARs) increased transiently at week 13 with 0.27 (0.18, 0.35; pBH=6.0e-9) while returning to baseline at week 26 (0.06; 0.03, 0.15; pBH=0.25). The sphingolipid classes of sphingomyelins (SMs), hexosylceramides (HexCers), dihexocylceramides (Hex2Cers) and trihexocylceramides (Hex3Cers) were increased at 13 weeks with 0.25 (0.17, 0.32; pBH=2.3e-10), 0.37 (0.13, 0.61; pBH=0.0055), 0.34 (0.16, 0.52; pBH=6.3e-4), 0.37 (0.07, 0.68; pBH=0.035), respectively, while the treatment effects were no longer statistically significant at week 26. In contrast to other sphingolipids, dihydroceramides (Cer(d)s) were unchanged at 13 weeks and decreased with -0.14 (-0.21, -0.08; pBH=7.0e-5) at 26 weeks. The other ten lipid classes, including cholesteryl esters (CEs), phosphatidylethanolamines (PEs) and phosphatidylserines (PSs), out of the 26 lipid classes measured remained unchanged throughout the trial.

### 3.4 The effect of liraglutide on molecular lipids

From all lipid classes, lysophospholipids showed the strongest positive association to treatment for both visits, with LPC(O)s showing a change for all measured lipids (p<0.05; Figure 2). The following LPC(O)s showed an effect (p<0.05) on both visits: LPC(O)s 16:0, 18:0, 20:0, 20:1, 22:0, 22:1, 24:0, 24:1, and 24:2. LPC(O-18:1) showed a statistically significant increase only at the 26-week time point. LPC(P)s 18:1 and 20:0 increased significantly on both visits, while LPC(P-18:0) only after 26 weeks. LPC 20:1 showed a statistically significant positive treatment effect on both visits, LPCs 19:1 and 26:0 only at 13-weeks, and LPCs 18:2, 20:0, and 22:0 only at 26 weeks. While LPEs showed a general trend of increase in plasma concentrations throughout the treatment, LPE 18:2 was the only LPE to show a statistically significant change.

**Figure 2.**
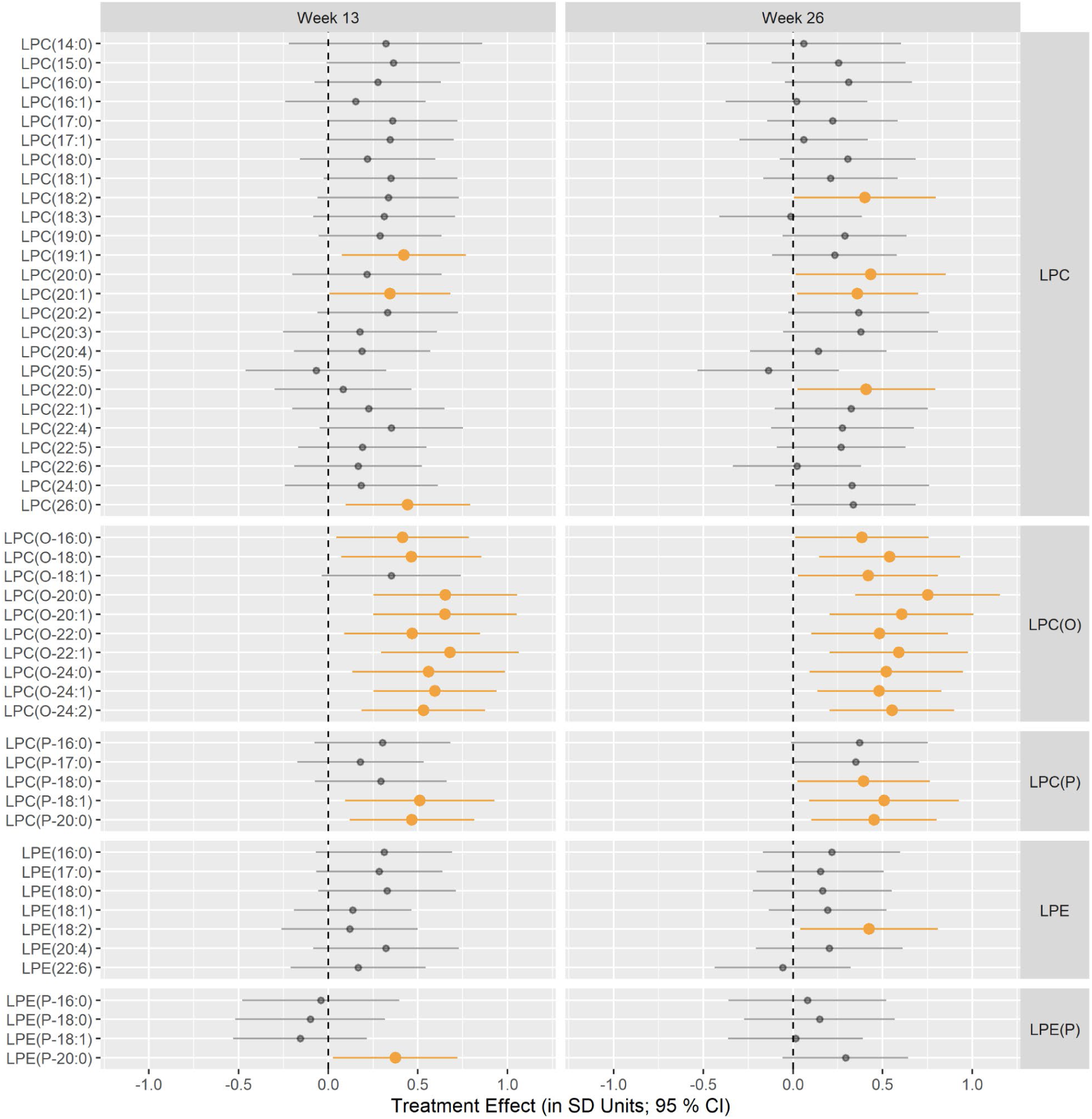
Treatment effect (x-axis) on molecular lysophospholipid species (y-axis) at 13 weeks (left) and 26 weeks (right). The results are categorized into panels according to lipid class (vertical division) and time point (horizontal panels). Treatment effects with p<0.05 are depicted in colour.

Treatment with liraglutide, resulted in reduced TG levels at 13 weeks, but this effect was not sustained at 26 weeks. FA levels remained at baseline concentrations at 13 weeks and declined at the end of treatment. The heavier TG species with a higher degree of unsaturation presented a stronger reduction throughout the trial (Figure 3). A mixed treatment effect was observed in PC(P) species, with both increases and decreases detected depending on the length of a carbon chain. The treatment effect according to lipid size and degree of unsaturation in all measured lipid classes is shown in Supplementary Figures 2 and 3.

**Figure 3.**
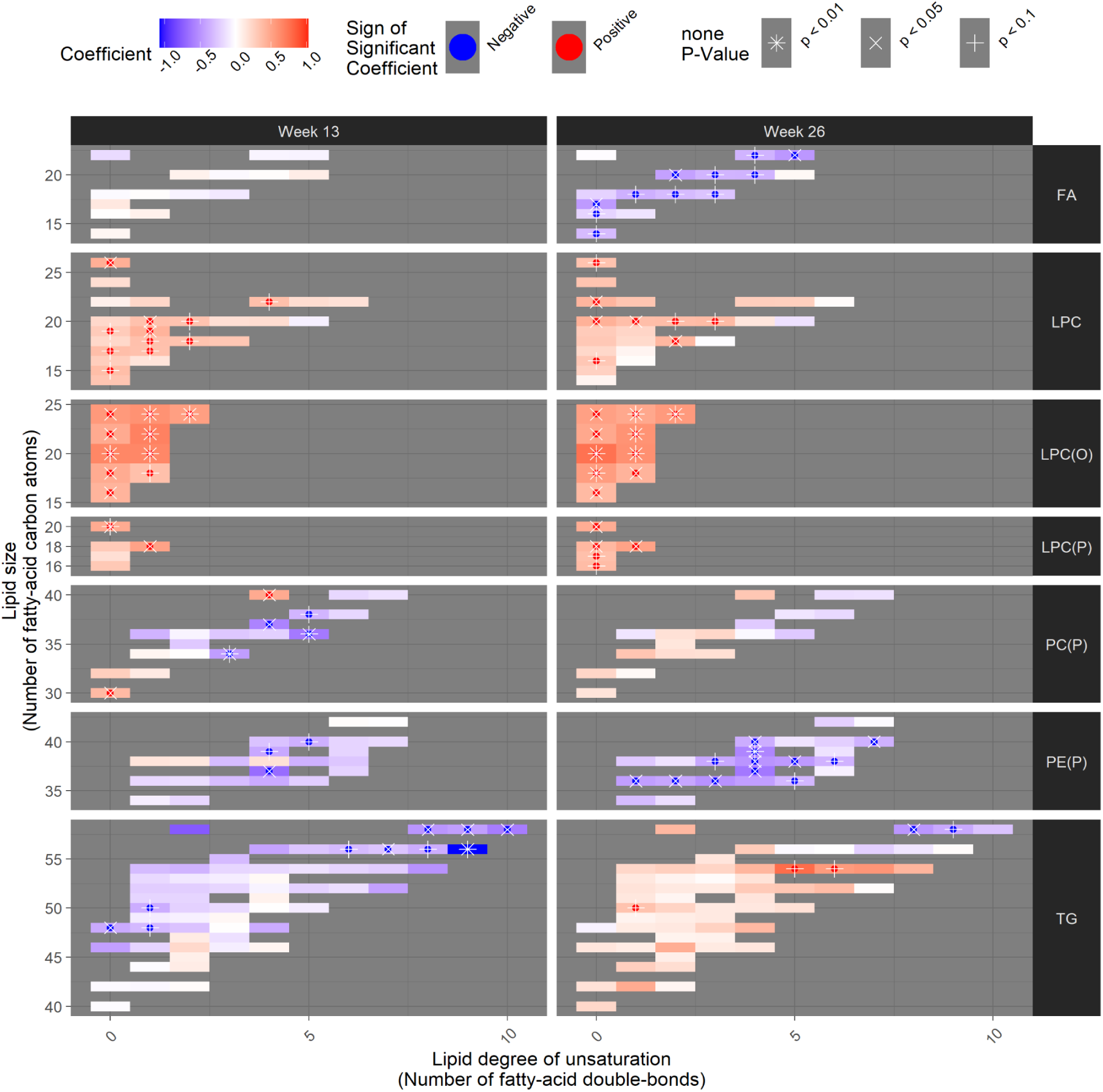
Treatment effect on the levels of molecular lipid species in classes with three or more affected lipid species (p<0.05). The results are categorized into panels according to lipid class (vertical division) and time point (horizontal division). Each coloured rectangle corresponds to one lipid species, where its position on the y-axis and x-axis, respectively, indicates its size and degree of unsaturation. The effect of the treatment is indicated by colour (red: increase; blue: decrease). The strength of the treatment effect is indicated with asterisk or cross for p<0.01 and p<0.05, respectively (Figure 1).

### 3.5 Associations between lipid changes and health biomarkers

We further investigated whether treatment-induced changes in lipid levels were associated with changes in clinical measurements that responded to the intervention, including the biomarkers of cardiac, hepatic, renal and immune function (Figure 4). Changes in lipid levels were independent of changes in BMI, except for the inverse association with Hex2Cers. Changes in HbA_1c_ were positively associated with changes in PEs and PE(O)s as well as in Cer(m)s, PC(O)s and SMs. Changes in ALAT were inversely associated with changes in the lysophospholipids LPCs, LPC(O)s, LPC(P)s, all of which increased during the trial, as well as with changes in PCs, OxPCs, OxLPCs, PC(P)s, PE(P)s and Cer(m)s. Changes in MCP-1 were inversely associated with changes in the phospholipids LPC(O)s, LPC(P)s, PC(O)s, LPE(P)s and in the sphingolipids SMs, Cer(d)s, Hex2Cers, and Hex3Cers. Finally, changes in UACR were inversely associated with changes in LPCs, LPC(O)s, and Cer(m)s, but positively with changes in OxPC levels.

**Figure 4.**
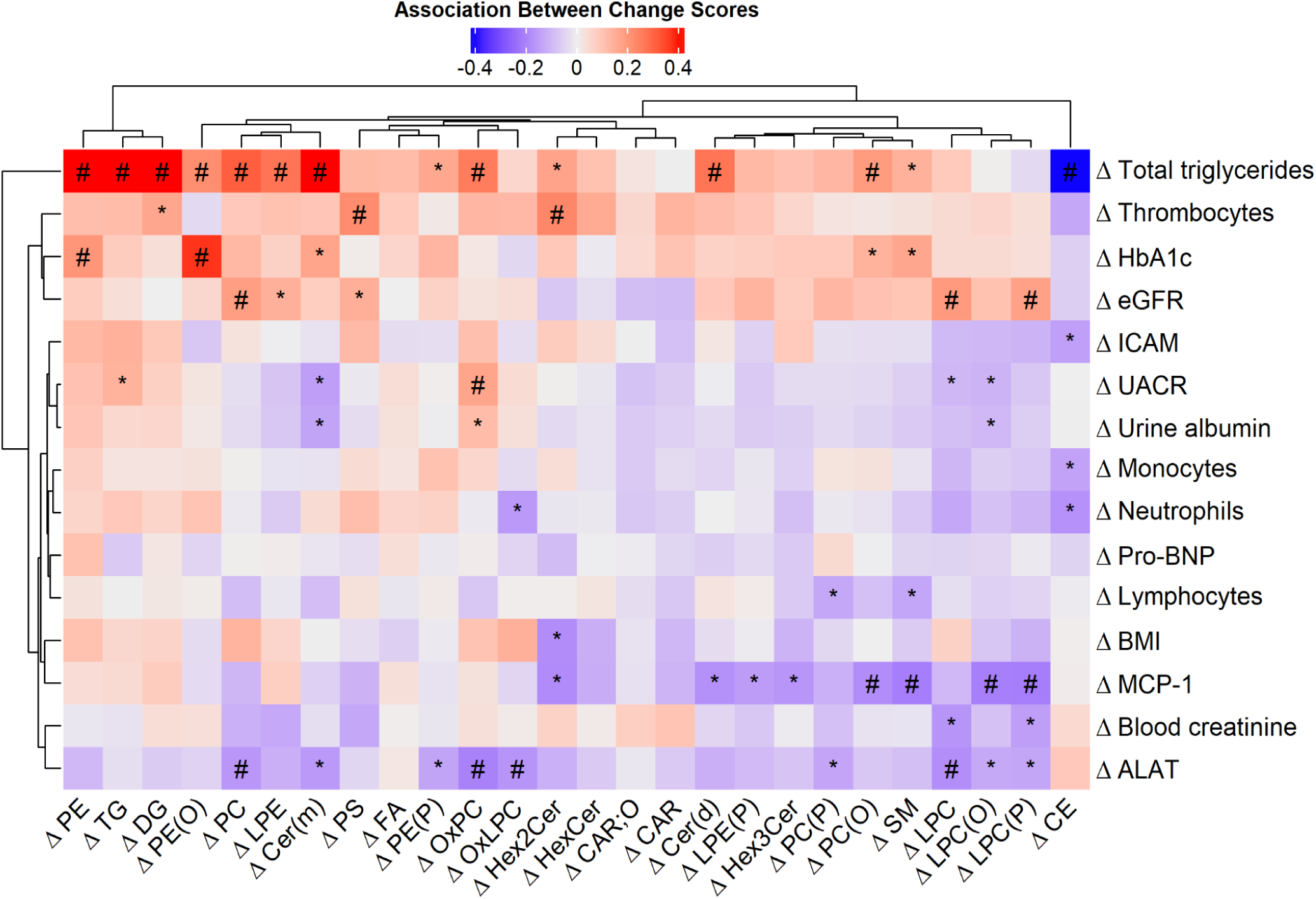
Association between the change in average level in lipid class (columns) and change in clinical measurements during the trial. Clinical measurements responding to treatment are included. Nominal p-values lower than 0.01 and 0.05 are highlighted with a sharp and an asterisk, respectively. (Example: LPC(O)s increased in the trial, and the change is inversely associated with changes in MCP-1 and ALAT, which both decreased with treatment).

The correlation between the lipids and clinical measurements was also investigated at baseline (Supplementary Figure 4). Glycerolipids as well as OxPCs and PEs were positively correlated with total triglycerides, UACR, and urine albumin (p<0.01). Additionally, ceramides (positively) and lysophospholipids (inversely) were correlated with total triglycerides. TGs, CARs and oxidised acylcarnitines (CAR;Os) were inversely correlated with eGFR. Lysophospholipids and Hex3Cers were inversely correlated with BMI, and LPC(O)s were inversely correlated with ALAT. Pro-BNP was not associated with lipid levels. Measurements related to the immune system were not correlated with lipid levels, except for MCP-1, which was positively correlated with FAs.

Finally, we investigated with mediation analysis whether the statistically significant treatment effects on clinical variables could be explained by the treatment effect on the most affected lipid class, the LPC(O)s. Here, the treatment effect on LPC(O) level did not mediate the treatment effect on BMI (average causal mediated effect, ACME, of 0.037 kg/m^2^ per SD-unit change in LPC(O); 95% CI: -0.15, 0.19; p=0.67). In reverse, the treatment effect on BMI also did not mediate the treatment effect on LPC(O) levels (ACME, of 0.034 SD-units per unit BMI change; 95% CI: -0.19, 0.28, p=0.75), indicating a direct treatment effect of liraglutide on the LPC(O) level instead of an indirect effect via BMI change. The treatment effects on other clinical variables that were affected by liraglutide (Figure 1A) were not mediated by the treatment effect on LPC(O) level, nor the reverse, with one exception: on thrombocytes 33% of the treatment effect (an increase) was mediated by the treatment effect on the LPC(O) level (ACME of 0.070 SD-units on thrombocytes per SD-unit increase in LPC(O); 0.0028, 0.18; p=0.036). There was no reverse mediation by thrombocytes’ treatment effect on the treatment effect of LPC(O) level (ACME of 0.039; -0.060, 0.17; p=0.46).

## 4. DISCUSSION

The time-dependent changes in lipid profiles observed with liraglutide treatment suggest dynamic lipid remodelling. The initial reduction in triglycerides at week 13, followed by fatty acid decline at week 26, is consistent with enhanced lipolysis and clearance through β-oxidation. The sustained increase in lysophospholipids may reflect enhanced phospholipid remodelling via the Lands cycle, in which phospholipase-mediated cleavage generates lysophospholipids that are then reacylated by lysophosphatidylcholine acyltransferase (LPCAT) enzymes using fatty acyl-CoA (Supplementary Figure 5).[27] These findings suggest a shift in the lipidome from a fatty acid-rich profile toward one enriched in lysophospholipids.

The treatment resulted in sustained reductions in BMI and HbA_1c_ throughout the 26-week trial. Additional improvements emerged at specific time points, including reductions in inflammatory markers MCP-1 and pro-BNP at week 13, followed by improvements in ALAT, ICAM, UACR at week 26. This temporal pattern suggests progressive cardiometabolic, hepatic and renal benefits. The observed decrease in eGFR may reflect hemodynamic changes in renal filtration associated with GLP-1RA treatment, although increased serum creatinine resulting from enhanced muscle catabolism during rapid weight loss may also contribute.

### 4.1 Liraglutide increases lysophospholipids

In the present study, liraglutide treatment significantly altered circulating lysophospholipid (LPC, LPC-O, LPC-P, LPE, LPE-P) levels in people with T2D. As a novel finding, we observed that plasma levels of all five subclasses increased as early as at 13 weeks and remained upregulated at week 26. Lysophospholipids are amphiphilic phospholipids with only one acyl chain produced from deacylation by the action of phospholipases.[28] Decreased levels of LPCs have been observed in people with obesity and T2D.[29–34] In a cross-sectional study, children with overweight and obesity exhibited reduced levels of LPCs, particularly LPC (0:0/16:0) and (16:0/0:0), while obesity management was associated with increased LPC and LPE levels.[35] Our longitudinal analysis demonstrated that liraglutide-induced increases in lysophospholipids were independent of the decrease in BMI. Mediation analysis further supported a direct effect of liraglutide on LPC levels rather than an indirect effect mediated through weight loss.

The literature suggests that LPCs exert both pro-inflammatory and anti-inflammatory effects[36,37], likely depending on the context. Research on lysophospholipids may yield different findings when investigated in cellular systems compared with blood samples as LPCs in cells are rapidly metabolised to lysophosphatidic acid.[37]

Various lysophospholipids have been studied in metabolic syndrome. In accordance with our results, Meikle et al. found decreased levels of LPC(O)s 22∶0, 24∶1, and 24∶2 in people with both T2D and prediabetes.[33] In our study, these lipids were significantly increased with liraglutide. Tonks et al.[29] found insulin resistance and/or obesity were associated with lower LPCs 20:0, 20:1, 22:0, 22:1, LPC(O)s 20:0, 22:1, 24:2, and LPE 16:0, which also have been significantly increased in our study. In the longitudinal METSIM study of Finnish men, lower baseline levels of LPCs, especially LPC 18:2, were identified as a predictor of incident T2D.[38] Consistent with its metabolic relevance, LPC 18:2 levels increased after 26 weeks of liraglutide treatment in the LIRAFLAME trial. It has been suggested that LPCs may play a protective role in regulating glucose homeostasis: especially LPCs 14:0 and 16:0 have the ability to bind to the G-protein coupled receptors GPR40, GPR50, and GPR119 on pancreatic β-cells, thus facilitating calcium transport, cyclic AMP (cAMP) synthesis, and glucose-stimulated insulin secretion.[39,40] Further studies should investigate the activity of LPCs and their role in the human body based on their structural differences.

### 4.2 Liraglutide reduces triglycerides and fatty acids

The effectiveness of GLP-1RA agents on lipid management in diabetes and obesity has been established in the literature. Liraglutide has been successful at regulating lipolysis, decreasing fat deposition and improving fat distribution from visceral to subcutaneous depots.[41] A review on GLP-1RA medications on lipid management summarised that these agents reduce circulating TGs and FAs[42], which was observed in our study. In the LIRAFLAME trial, FAs showed a significant reduction at 26 weeks of liraglutide treatment, while TGs were decreased at 13 weeks before returning towards baseline at 26 weeks. These findings suggest improved lipid handling associated with significant improvements in BMI, HbA_1c_ and cholesterol. Lowering of plasma FAs is beneficial, as increased serum levels contribute to insulin resistance (IR) and cardiovascular diseases.[43]

The metabolism of TGs into FAs is a complex mechanism involving various enzymatic lipolysis and proteins whose action may be influenced by a GLP1-RA agent. In a recent study investigating proteomic changes in people with obesity, semaglutide was found to significantly increase pancreatic lipase (PNL) and two assisting proteins, pancreatic lipase-related proteins PNLRP1 and PNLRP2.[44] Given that pancreatic lipase hydrolyses 50–70% of dietary triglycerides[45], its upregulation by liraglutide may contribute to the reduction in TGs observed in our study at the 13-week point. The decrease is consistent with more efficient lipoprotein lipase (LPL)-mediated lipid breakdown and improved triglyceride metabolism, as liraglutide has been reported to increase serum lipase by 28% after 6 months of treatment.[46]

In the LEAD trial, including over 1000 individuals with T2D treated with liraglutide for 26 weeks, a significant reduction in TG levels was reported.[47] The GLP1-RA has also been reported to decrease both fasting and postprandial levels of apolipoprotein C-III, which slows down the breakdown of TGs by inhibiting LPL.[48] Liraglutide has been shown to reduce postprandial apoB48 concentrations, which decreases dyslipidaemia and cardiovascular risk.[49] A plausible mechanism underlying the reduction in circulating triglycerides involves suppression of intestinal chylomicron production.[50,51] Following nutrient ingestion, GLP-1 receptor activation is thought to initiate a gut–brain–gut neural circuit via vagal afferent signalling, ultimately reducing the secretion of triglyceride-rich chylomicrons from enterocytes.[50,51] This effect is associated with reduced expression of key proteins involved in chylomicron assembly, including apolipoprotein B48 (ApoB48), microsomal triglyceride transfer protein (MTP), and diacylglycerol acyltransferase-1 (DGAT1). ApoB48 provides the structural scaffold for chylomicron formation, MTP transfers triglycerides onto ApoB48 during lipoprotein assembly, and DGAT1 is the enzyme responsible for the final step of triglyceride synthesis.[52,53] Together, these changes impair chylomicron assembly and reduce postprandial triglyceride secretion. Although early studies suggested a direct regulation of enterocyte lipoprotein production by GLP-1RAs[54], current evidence indicates that this process is mediated predominantly through gut–brain neural signalling, while a direct intestinal effect cannot be excluded.[50,51] Further research linking the lipidome to proteome is needed to understand the underlying mechanisms of liraglutide’s effect on TG metabolism (see proposed mechanism in Supplementary Figure 5).

### 4.3 Liraglutide effect on lysophospholipids is independent of weight loss

In this study, liraglutide was effective in reducing body weight, and HbA_1c_, which are broadly associated with inflammation.[55] The treatment was effective at improving biomarkers relevant to cardiovascular, hepatic, and renal health, including ALAT, MCP- 1, ICAM, pro-BNP, and urinary albumin. The correlation between the upregulated LPC(O)s and decreases in MCP-1, ALAT and UACR suggests a link between anti-inflammatory, renal and liver health benefits and this lipid family. ALAT is a liver injury biomarker, usually elevated in people with T2D.[56] The correlation of the decrease in ALAT with the increase in LPC(P)s, LPC(O)s, and LPEs demonstrated that liraglutide may be linked to improvement in liver health.

In addition, elevated LPC(O/P)s were associated with reduced MCP-1 levels, a key inflammatory chemokine involved in monocyte recruitment and linked to both kidney disease progression and cardiovascular risk.[57] This inverse relationship may point to a potential anti-inflammatory role of specific LPC species. Moreover, plasmalogen-type LPCs have been described to protect membranes from oxidative stress.[58]

The LPC(O) levels increased throughout the trial. This treatment effect mediated one-third of the increase in thrombocyte levels, suggesting that LPC(O)s are involved in platelet biology.[59] Mediation analysis confirmed that the remodelling of LPC(O) species by liraglutide represents a direct treatment effect, independent of changes in BMI. Neither BMI nor any other liraglutide-affected clinical variable mediated the LPC(O) response, indicating that remodelling is not a downstream consequence of weight loss but a primary pharmacological effect of GLP-1 receptor agonism.

## 5. Strengths and limitations

A key strength of this study is its double-blind, randomised, placebo-controlled design, with targeted lipidomic profiling performed at three distinct time points. This approach enabled assessment of temporal changes in lipid metabolism during treatment and further mediation analyses to infer causal links. However, the study duration was relatively short, and future investigations with extended follow-up periods, including annual assessments, should be conducted. Several limitations should also be acknowledged. As the lipidomic analysis was carried out post-hoc, the results should be validated in an external cohort. Although treatment effects were estimated using models adjusted for total TG levels, the observed baseline imbalance in TG concentrations between the groups should be considered a limitation as the residual effects of the baseline TG imbalance cannot be ruled out. In addition, the study population included a relatively low proportion of female participants, potentially restricting the ability to generalise the findings across sexes. All participants were of White ethnicity, which may limit the generalisability of the results across different ethnic groups.

## 6. Conclusions

We observed a consistent upregulation of lysophospholipids, with the strongest increase in lysoalkylphosphatidylcholines (LPC(O)s), at 13 weeks of liraglutide treatment that was sustained at 26 weeks. The change was inversely associated with ALAT, MCP-1, and UACR, suggesting potential links to anti-inflammatory mechanisms with potential hepatic, renal, and cardiovascular benefits. The temporal pattern of the triglyceride reduction at 13 weeks followed by fatty acid decline at 26 weeks is consistent with lipid remodelling. Lysophospholipids responding to liraglutide warrant further investigation to assess the causal links to health outcomes.

## Data Availability

All data produced in the present study are available upon reasonable request to the authors

## Author Contributions

The author contributions are as follows: study concept and design (PR, CLQ); acquisition of data (SMK, TS); analysis and interpretation of data (TS, DL, CLQ); original study (PR, RSR, EZ); drafting the first version of the manuscript (DL, TS, CLQ); critical revision of the manuscript (all authors).

## Funding

The lipidomics analysis was funded by Novo Nordisk A/S. LIRAFLAME was funded by Novo Nordisk A/S and Skibsreder Per Henriksen, R. Oghustrus fund. Steno Diabetes Center Copenhagen and the Department of Clinical Physiology, Nuclear Medicine & PET, Rigshospitalet & Cluster for Molecular Imaging, University of Copenhagen, Denmark, have provided internal funding (ERC Advanced Grant no. 670261; Research Foundation of Rigshospitalet; Research Council of the Capital Region of Denmark; Lundbeck Foundation; Novo Nordisk Foundation; The John and Birthe Meyer Foundation).

## Ethical Approval

The studies involving human participants were reviewed and approved by the regional ethics committee for RegionH (H-16044546) and the Danish Medicines Agency (2016110109) and were performed in compliance with the principles of the Declaration of Helsinki. The patients/participants provided their written informed consent to participate in this study.

## Availability of Data and Materials

The dataset analysed here is not publicly available, for the privacy of the participants, in compliance with EU and Danish data protection law. The data can be accessed upon reasonable request; relevant legal permission from the data protection agency is required. Data access requests should be directed to PR.

## Disclosure Statement

BJS, EHZ, MAR, KS and JMLM are present employees of Novo Nordisk A/S. Other authors declare no conflict of interest associated with their contribution to this manuscript. CLQ has received consultancy fees from Pfizer, honoraria, travel or speakers’ fees from Biogen, and research funds from Pfizer, Novo Nordisk and Waters Corporation. CLQ is director and founder of the consultancy company BrainLogia. PR has received grants from Astra Zeneca, Bayer, Novo Nordisk and Lexicon Pharma, and honoraria to Steno Diabetes Center Copenhagen from Amgen, Astra Zeneca, Abbott, Bayer, Boehringer Ingelheim, Lexicon, Novo Nordisk, Roche, Regeneron. RSR has received consultancy fees from Novo Nordisk A/S. TWH and RSR own shares in Novo Nordisk A/S.

## Abbreviations

T2D: Type 2 Diabetes
GLP-1RA: Glucagon-Like Peptide-1 Receptor Agonist
HbA_1c_: Glycated Haemoglobin A_1c_
BMI: Body Mass Index
eGFR: Estimated Glomerular Filtration Rate
ALAT: Alanine Aminotransferase
MCP-1: Monocyte Chemoattractant Protein-1
LPC: Lysophosphatidylcholine
TG: Triglyceride
LPC(O): Lysoalkylphosphatidylcholine
LPC(P): Lysoalkenylphosphatidylcholine
LPE: Lysophosphatidylethanolamine
FA: Fatty Acid
pBH: Benjamini-Hochberg-corrected p- value
ACME: Average Causal Mediated Effect, urinary albumin-to-creatinine ratio (UACR).

## Notes

### Clinical Trial

NCT03449654

