## Supplementary Materials for "Glucagon-like peptide-1 receptor agonist-induced lipidome remodelling is associated with improved liver, kidney and inflammatory markers in type 2 diabetes"

**Supplementary Table 1.** Treatment effect on clinical characteristics at weeks 13 and 26.

| Variable | Treatment Effect at Week 13 | P-Value at Week 13 | Treatment Effect at Week 26 | P-Value at Week 26 |
|---|---|---|---|---|
| **BMI (kg/m$^2$)** | -1.01 (-1.25, -0.77) | 2.4e-15 | -1.04 (-1.28, -0.80) | 3.1e-16 |
| **$HbA_{1c}$ (mmol/mol)** | -6.59 (-8.75, -4.44) | 4.1e-09 | -4.90 (-7.06, -2.73) | 1.1e-05 |
| **HDL cholesterol (mmol/L)** | -0.05 (-0.10, 0.01) | 0.10 | -0.00 (-0.06, 0.06) | 0.95 |
| **LDL cholesterol (mmol/L)** | 0.01 (-0.19, 0.21) | 0.89 | 0.15 (-0.05, 0.35) | 0.15 |
| **VLDL cholesterol (mmol/L)** | -0.04 (-0.13, 0.05) | 0.35 | -0.06 (-0.15, 0.03) | 0.18 |
| **Total cholesterol (mmol/L)** | -0.14 (-0.36, 0.07) | 0.20 | 0.09 (-0.13, 0.32) | 0.41 |
| **Total triglycerides (mmol/L)** | -0.30 (-0.55, -0.05) | 0.019 | 0.01 (-0.24, 0.26) | 0.91 |
| **Alanine aminotransferase (IU/L)** | -2.62 (-5.90, 0.66) | 0.12 | -7.17 (-10.47, -3.87) | 2.4e-05 |

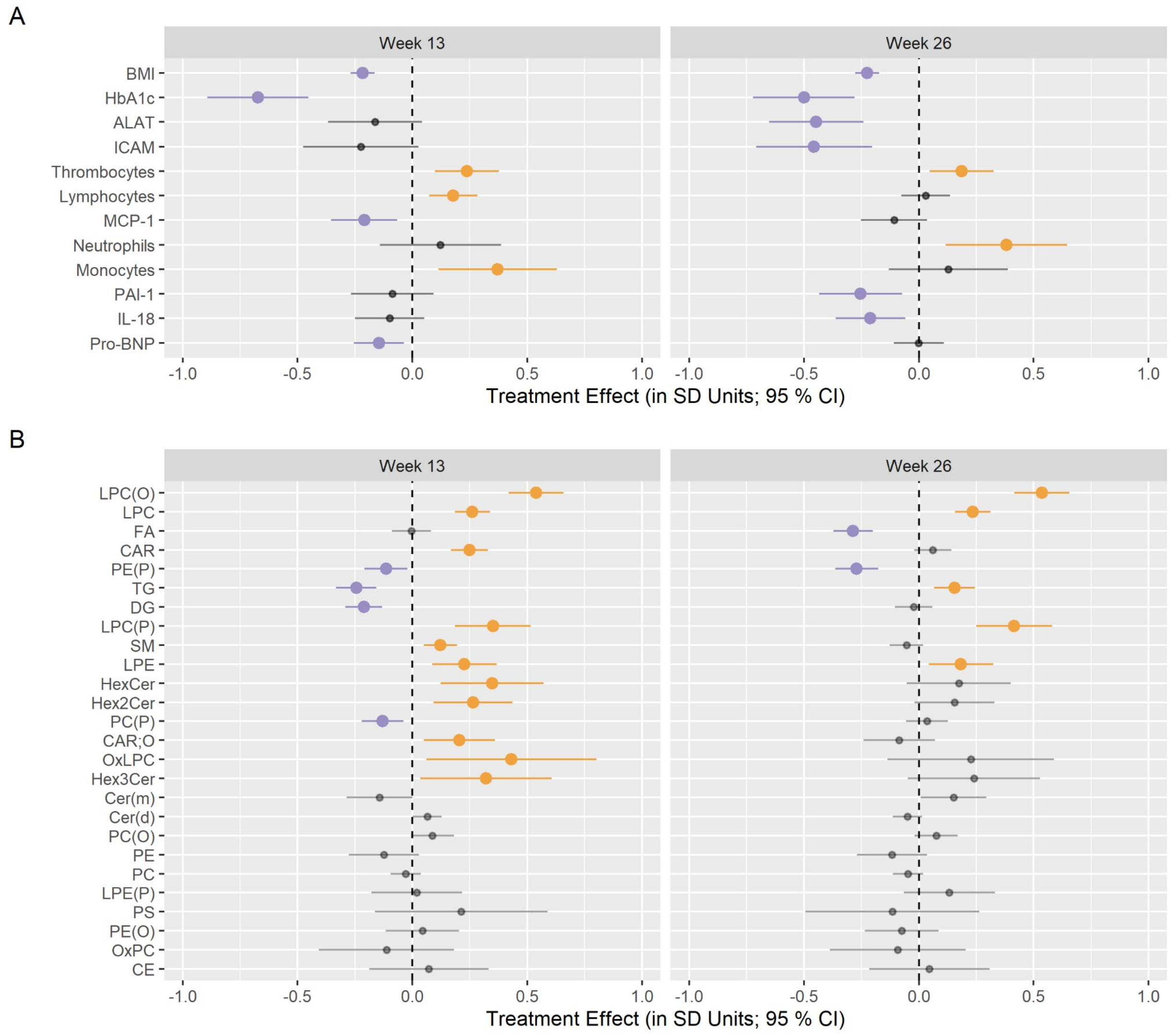


**Supplementary Figure 1.** The effect of liraglutide treatment on the levels in A) clinical variables and B) lipid classes after 13 weeks (left) and 26 weeks (right) of treatment, calculated without adjustment for the baseline values of covariates. Increases and decreases with statistical significance (pBH<0.05) are highlighted in orange and purple, respectively.

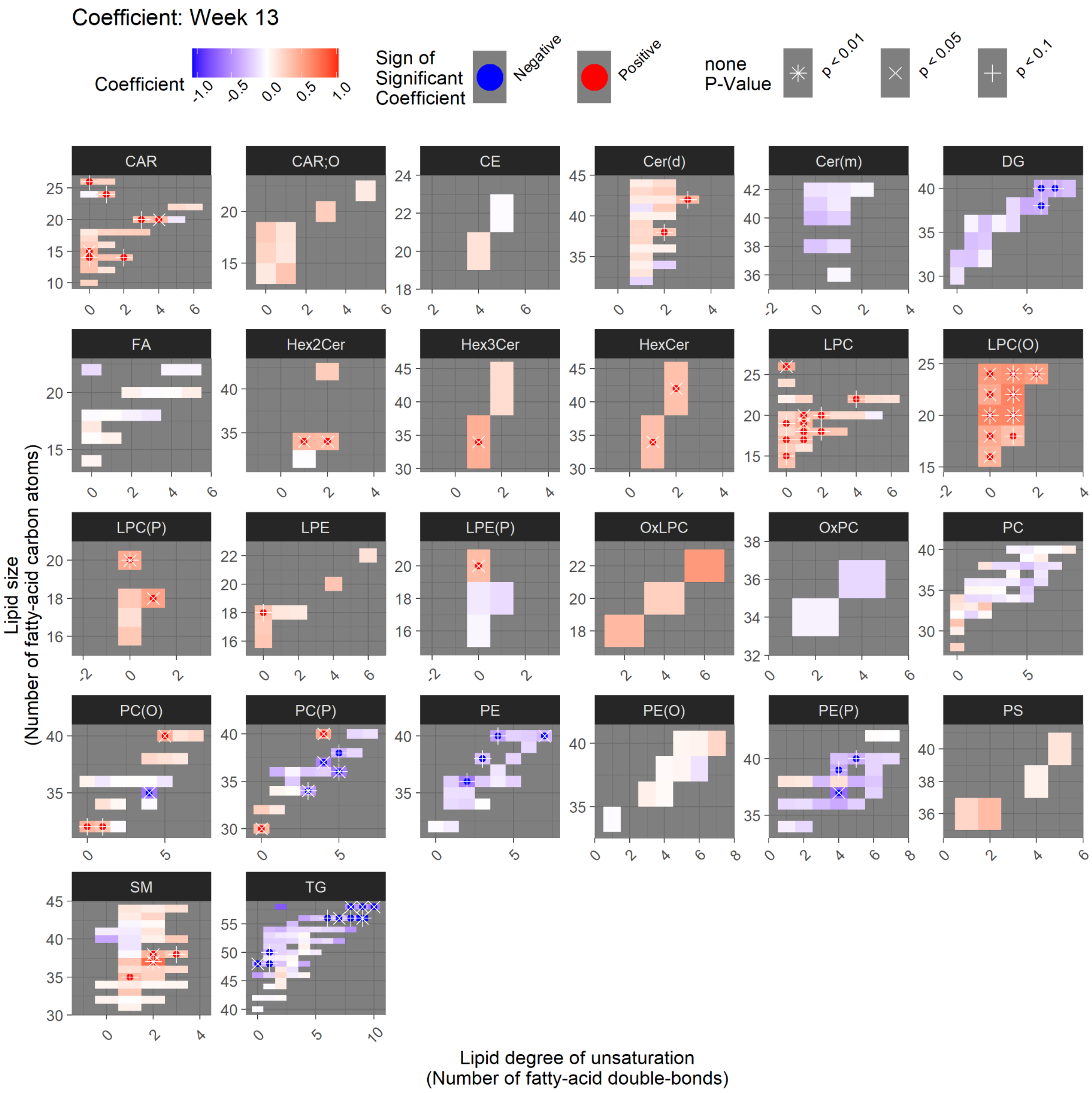


**Supplementary Figure 2:** Treatment effect on the levels of molecular lipid species at week 13. The lipid species are categorized into panels according to lipid class. Each coloured rectangle corresponds to one lipid species, where its position on the y-axis and x-axis, respectively, indicates its size and degree of unsaturation. The effect of the treatment is indicated by colour (red: increase; blue: decrease). The strength of the treatment effect is indicated with an asterisk or cross for p<0.01 and p<0.05, respectively.

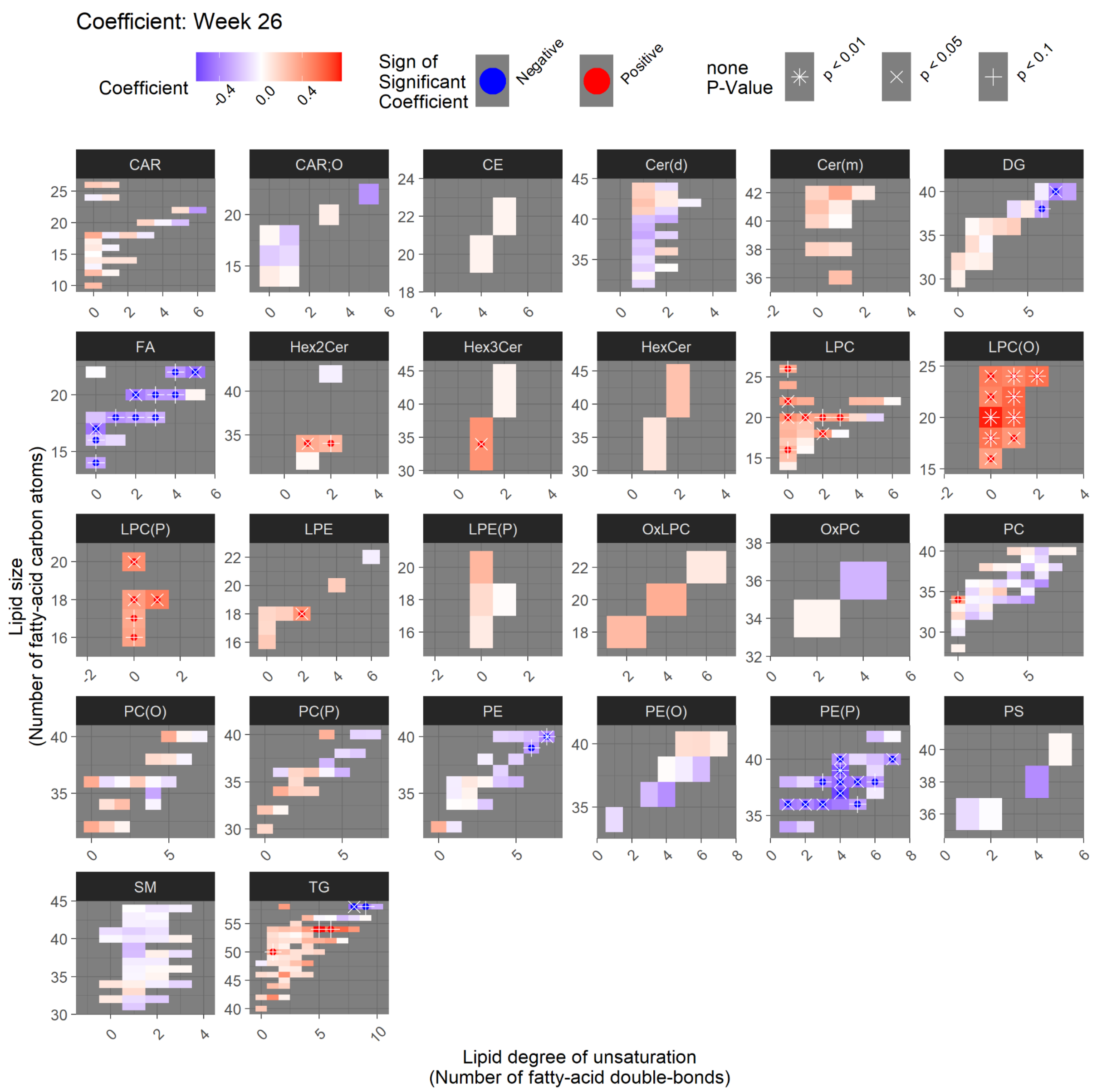


**Supplementary Figure 3:** Treatment effect on the levels of molecular lipid species at week 26. The lipid species are categorized into panels according to lipid class. Each coloured rectangle corresponds to one lipid species, where its position on the y-axis and x-axis, respectively, indicates its size and degree of unsaturation. The effect of the treatment is indicated by colour (red: increase; blue: decrease). The strength of the treatment effect is indicated with an asterisk or cross for $p<0.01$ and $p<0.05$, respectively.

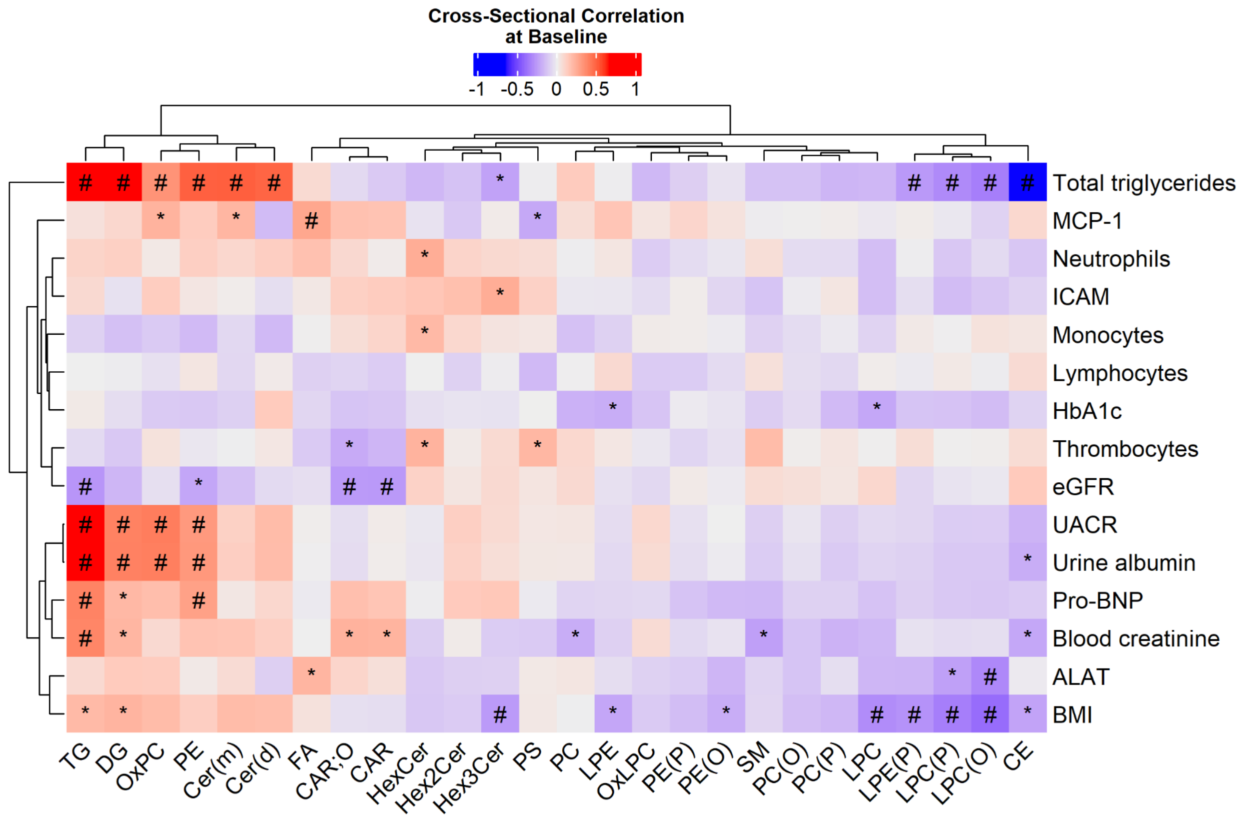


**Supplementary Figure 4:** Cross-sectional correlation at the baseline visit between average level in lipid class (columns) and clinical measurements (rows). Clinical measurements responding to treatment are included. Nominal p-values smaller than 0.01 and 0.05 are highlighted with a sharp and an asterisk, respectively.

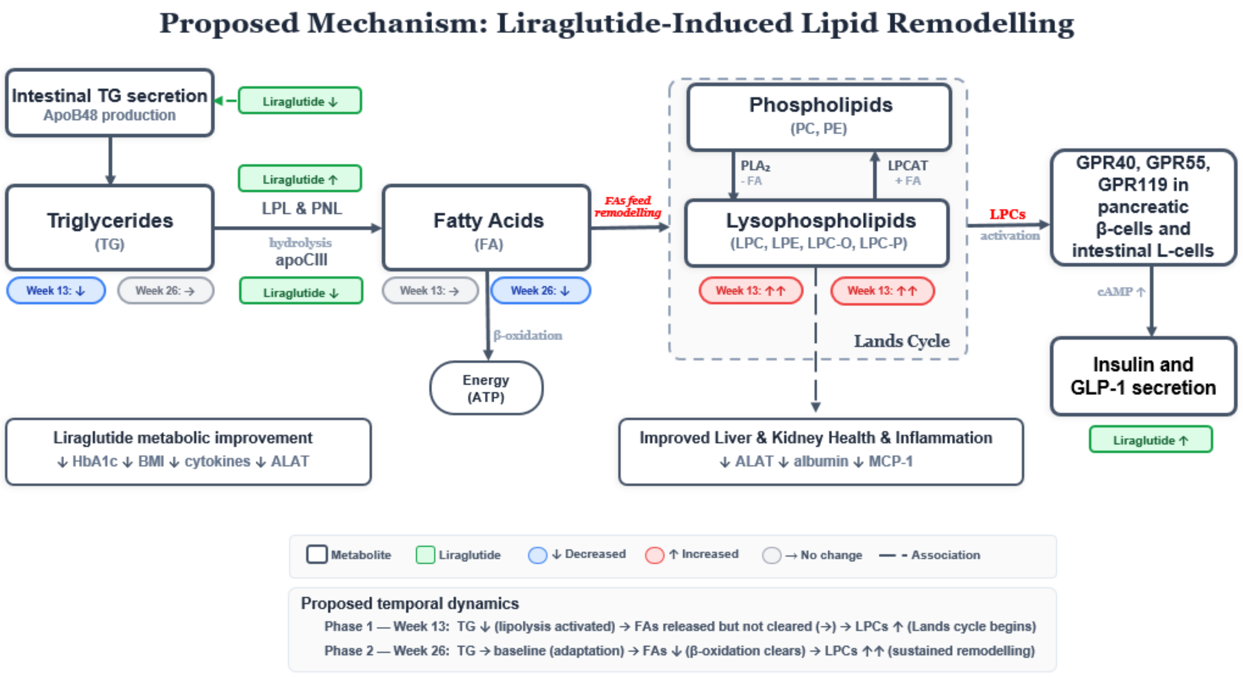


**Supplementary Figure 5:** Proposed mechanism of liraglutide-induced lipid remodelling.
